# Possible involvement of keratinocyte-derived microvesicle particles in human photosensitivity disorders

**DOI:** 10.1101/2025.09.24.25336388

**Authors:** Risha Annamraju, Madison S. Owens, Anita Thyagarajan, Danielle A. Corbin, Catherine M.T. Sherwin, Jade Bryant, Garrett W. Fisher, Winston R. Owens, Alycia Ketter, Craig A. Rohan, Michael G. Kemp, Robyn K. Crow, Jeffrey B. Travers

**Author notes:** Corresponding author: Jeffrey B. Travers, M.D., Ph.D. Wright State University Department of Pharmacology & Toxicology, 3640 Colonel Glenn Hwy, Dayton OH. These two authors contributed equally to this manuscript (Risha Annamraju, Madison S. Owens).

## Abstract

**Background:** Previous murine studies have implicated acid sphingomyelinase-(aSMase) generated subcellular microvesicle particles (MVP) in photosensitivity. Objective: The current double-blinded placebo-controlled studies examined if a single localized ultraviolet B radiation (UVB) treatment generated more MVP in human subjects with self-identified photosensitivity versus normal controls. A topical 4% formulation of the aSMase inhibitor imipramine applied immediately after UVB blocked the MVP release and erythema responses. Erythema responses at 24 and 72 h in response to multiple UVB fluences and minimal erythema doses (MED) at 24 h and effects of imipramine were also tested.

**Results:** Small cohorts of 10 adult self-identified photosensitive subjects and 10 controls were enrolled in these pilot studies which revealed increased levels of skin MVP in UVB-treated photosensitive subjects over controls which correlated with MED values. Moreover, post-UVB application of imipramine blunted UVB-induced MVP responses as well as tended to diminish erythema levels at 4 h but not at 24- or 72 h in photosensitive patients.

**Conclusion:** Though limited by low numbers of self-identified subjects, these pilot studies provide some support for the hypothesis that MVP could be involved in multiple types of human photosensitivity responses and suggest aSMase inhibition as a potential therapeutic strategy.

## Introduction

Photosensitivity is defined as an extreme response to ultraviolet radiation (UVR) from the sun or other sources and can be connected with many underlying diseases (Foering et al., 2013, Kim and Chong, 2013; Lehmann and Schwarz, 2011). Photosensitivity treatments revolve around limiting UVR exposure as well as strategies to block its absorption and effects. There are significant knowledge gaps as to how photosensitivity occurs which has resulted in therapeutic deficits.

Recent reports have implicated the release of keratinocyte subcellular microvesicle particles (100 nm-1000 nm; MVP; large extracellular vesicles) in response to the glycerophosphocholine-derived lipid mediator Platelet-activating factor (1-alkyl-2-acetyl glycerophosphocholines; PAF) in ultraviolet B radiation (UVB) signaling (Frommeyer et al., 2022). PAF acts on a single G-protein coupled receptor (PAFR) expressed on multiple cell types including granulocytes, B cells and epithelial cells such as the keratinocyte (Travers, 2020). Though the most potent lipid mediator yet described, PAF’s effects are limited as this family of lipids are metabolically labile, being quickly metabolized to biologically inert lyso-glycerophosphocholines by serum- and cell-associated acetyl hydrolases (McIntyre et al., 2009). In the context of UVB, preclinical studies have demonstrated that PAFR signaling is involved in acute processes such as inflammation and nociception (Zhang et al., 2009). The PAF system is also involved UVB-induced systemic immunosuppression in a process involving the mast cell PAFR with effector regulatory T cells (Bernard et al., 2019; Liu et al., 2021). Both murine and human studies have indicated that PAF travels in MVP which are hypothesized to protect this labile glycerophosphocholine from degradation (Liu et al., 2021; Lohade et al., 2024).

Preclinical studies using the xeroderma pigmentosum type A (XPA) protein deficiency photosensitivity model has indicated that PAFR-signaling and the MVP-generating enzyme acid sphingomyelinase (aSMase) are involved with the increased UVB-but not phorbol ester-induced skin inflammation in XPA deficient mice (Yao et al., 2012; Christian et al., 2024). Moreover, a recent report demonstrated that UVB-treatment of the NZM2328 mouse, a model for lupus erythematosus, also resulted in increased skin and systemic MVPs as compared to control mice (Corbin et al., 2023). These preclinical findings are suggestive that multiple types of photosensitivity could involve PAF and MVP.

The goal of the present pilot studies is to test the hypothesis that human subjects with photosensitivity respond to localized UVB irradiation with increased skin MVP, and test the ability of the functional inhibitor of aSMase (FIASM) imipramine (Beckmann et al., 2014; Christian et al., 2024; Lohade et al., 2024) to attenuate MVP production. We also tested if the UVB-induced erythema at both early and later time points can also be affected by post-UVB treatment of topical impramine.

## Results and Discussion

The goal of our IRB-approved pilot studies was to ascertain if a localized UVB treatment to skin resulted in differences in cutaneous MVP between subjects with self-identified photosensitivity versus control subjects. See CONSORT diagram in Figure 1a. Eleven subjects in each cohort were screened. One control subject screen failed as they were taking doxycycline, a known photosensitive drug. One of the photosensitive subjects was found after finishing the protocol to have been taking duloxetine, a known FIASM (Kornhuber et al., 2008). Thus, we excluded one photosensitive patient and one control subject, and performed this study with a cohort of ten patients and ten controls. Our acute 4 h protocol (Figure 1b) was somewhat similar to that previously published using control subjects consisting of young (18–35-year-old) males which revealed that high dose of oral antioxidant vitamins blocked UVB-generated MVP (McClone et al., 2022). The subjects with clinical photosensitivity and controls were recruited from Wright State University Department of Dermatology clinics. As shown in Table I, many of the self-identified photosensitive subjects carried diagnoses including lupus erythematosus and dermatomyositis. Of importance, these subjects continued their current immunomodulatory medications including hydroxychloroquine.

**Table 1.** Subject Characteristics and Data. Subject characteristics including age, sex, Fitzpatrick Skin type, diagnoses, relevant therapeutics and data. Abbreviations. Photo: Photosensitive subject; DLE: Discoid lupus erythematosus; DM: dermatomyositis; UNK; Unknown disorder; PMLE: polymorphous light eruption; HCQ: Hydroxychloroquine; MMF: Mycophenolate mofetil; MTX: Methotrexate; MED: Minimal erythema dose on back skin at 24 h post-UVB.

| Subject Type | Age (decade) /skin type | Diagnosis | Medications | Fold UVB+VEH MVP | Fold UVB+I MIP MVP | Fold UVB+VEH Erythema | Fold UVB+IMIP Erythema | MED 24h (J/m<sup>2</sup>) |
| --- | --- | --- | --- | --- | --- | --- | --- | --- |
| Control | 40s IV |  |  | 0.19 | 0.18 | 1.14 | 1.42 | 600 |
| Control | 30s III |  |  | 2.44 | 0.47 | 1.90 | 2.04 | 100 |
| Control | 20s III |  |  | 0.06 | 0.76 | 1.64 | 1.57 | 200 |
| Control | 30s I |  |  | 0.95 | 1.71 | 2.69 | 2.85 | 200 |
| Control | 20s I |  |  | 1.79 | 0.53 | 1.51 | 1.38 | 400 |
| Control | 60s II |  |  | 0.81 | 0.29 | 1.43 | 1.56 | 400 |
| Control | 30s II |  |  | 0.86 | 1.00 | 2.12 | 2.10 | 200 |
| Control | 20s II |  |  | 3.35 | 1.98 | 1.53 | 1.40 | 100 |
| Control | 20s II |  |  | 1.03 | 1.23 | 1.66 | 0.84 | 400 |
| Control | 70s II |  |  | 0.75 | 1.20 | 1.71 | 1.65 | 400 |
| **AVERAGE (SD)** | **37.1 (18)** |  |  | **1.22 (1.02)** | **0.94 (0.61)** | **1.73 (0.43)** | **1.68 (0.52)** |  |
| Photo | 50s II | DLE |  | 2.87 | 1.03 | 1.67 | 1.08 | 100 |
| Photo | 70s II | DLE | HCQ | 2.17 | 1.29 | 1.89 | 1.65 | 200 |
| Photo | 80s II | DM | MMF | 5.02 | 1.36 | 1.13 | 0.83 | 200 |
| Photo | 50s V | DLE | MTX, Prednisone | 2.80 | 0.55 | 1.00 | 1.07 | 400 |
| Photo | 60s I | DM | HCQ, MMF | 1.74 | 1.58 | 1.02 | 1.11 | 400 |
| Photo | 50s I | DM | MMF | 2.17 | 0.91 | 1.45 | 0.63 | 200 |
| Photo | 40s I | UNK | Pregabalin | 1.74 | 0.88 | 1.23 | 1.04 | 200 |
| Photo | 30s V | UNK | HCQ | 4.00 | 1.84 | 0.87 | 0.91 | 100 |
| Photo | 60s I | UNK | HCQ | 2.86 | 1.43 | 1.10 | 1.16 | 100 |
| Photo | 50s I | PMLE |  | 3.02 | 1.61 | 1.37 | 0.97 | 200 |
| **AVERAGE Age (SD)** | **59 (14)** |  |  | **2.84 (1.02)** | **1.25 (0.40)** | **1.27 (0.32)** | **1.04 (0.26)** |  |
| Excluded |  |  |  |  |  |  |  |  |
| Photo | 50s II | DM | Duloxetine | 0.75 | 1.10 | 0.97 | 1.12 | 400 |

**Figure 1.**
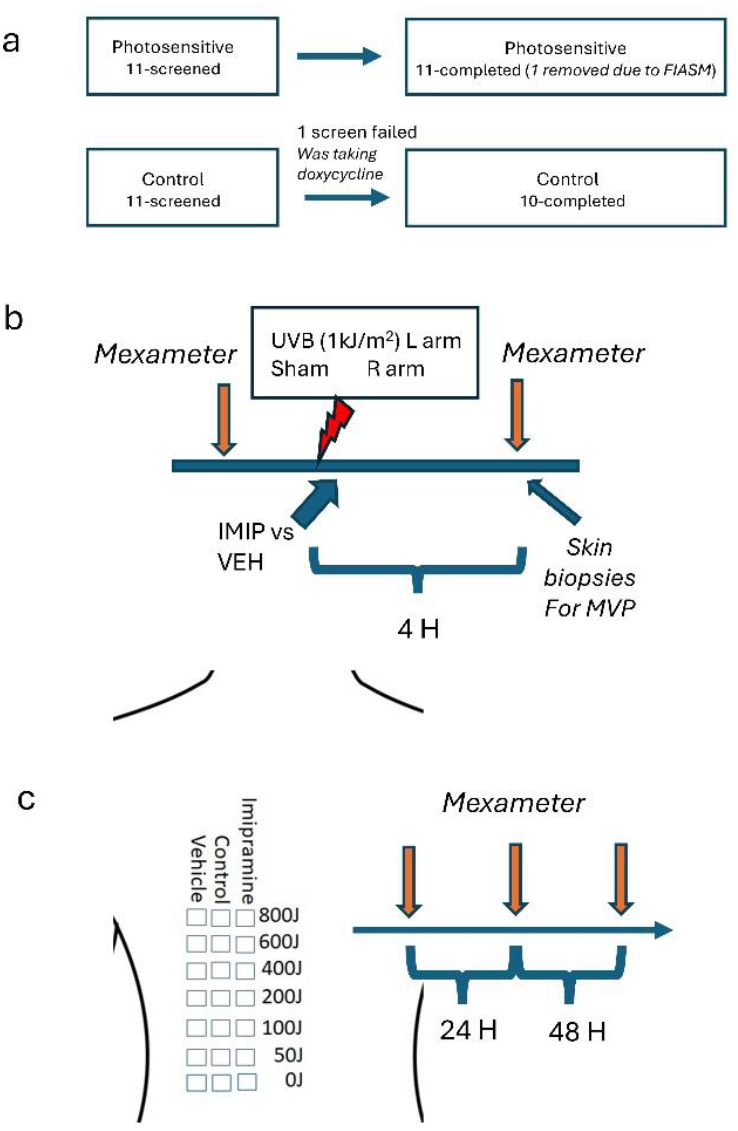
CONSORT Diagram and clinical trial protocols. **a)** CONSORT diagram. Please note one subject in the control group screen-failed as was taking the photosensitizing antibiotic doxycycline, and one photosensitive subject finished the study but was found after completion to have been taking a known FIASM. **b, c)**. Clinical trial protocols for the **b)** 4h forearm and **c)** 24,72h back studies.

The levels of MVP following a single UVB treatment to a 1 x 1 cm2 area on volar forearms were increased in subjects who were photosensitive (Figure 2c,d) versus control subjects (Figure 2a,b). To evaluate the role of aSMase in both MVP release as well as the UVB-induced erythema, we used imipramine, a known inhibitor of aSMase. There are a large group of molecules that have been demonstrated to serve as FIASM (Kornhuber et al., 2008; Kornhuber et al., 2010). Studies suggest that these compounds which are weak bases, can act in lysosomes to disrupt aSMase binding and thus accelerates the metabolism of this important MVP-generating enzyme. Topical application of 4% imipramine immediately following the UVB treatment inhibited the MVP levels in skin in the photosensitive subjects but not the control subjects (Please see Table I with all data from the individual subjects). It should be noted that imipramine was added immediately following the UVB to ensure that this compound did not exert a “sunscreen” effect through direct absorption of UV wavelengths.

**Figure 2.**
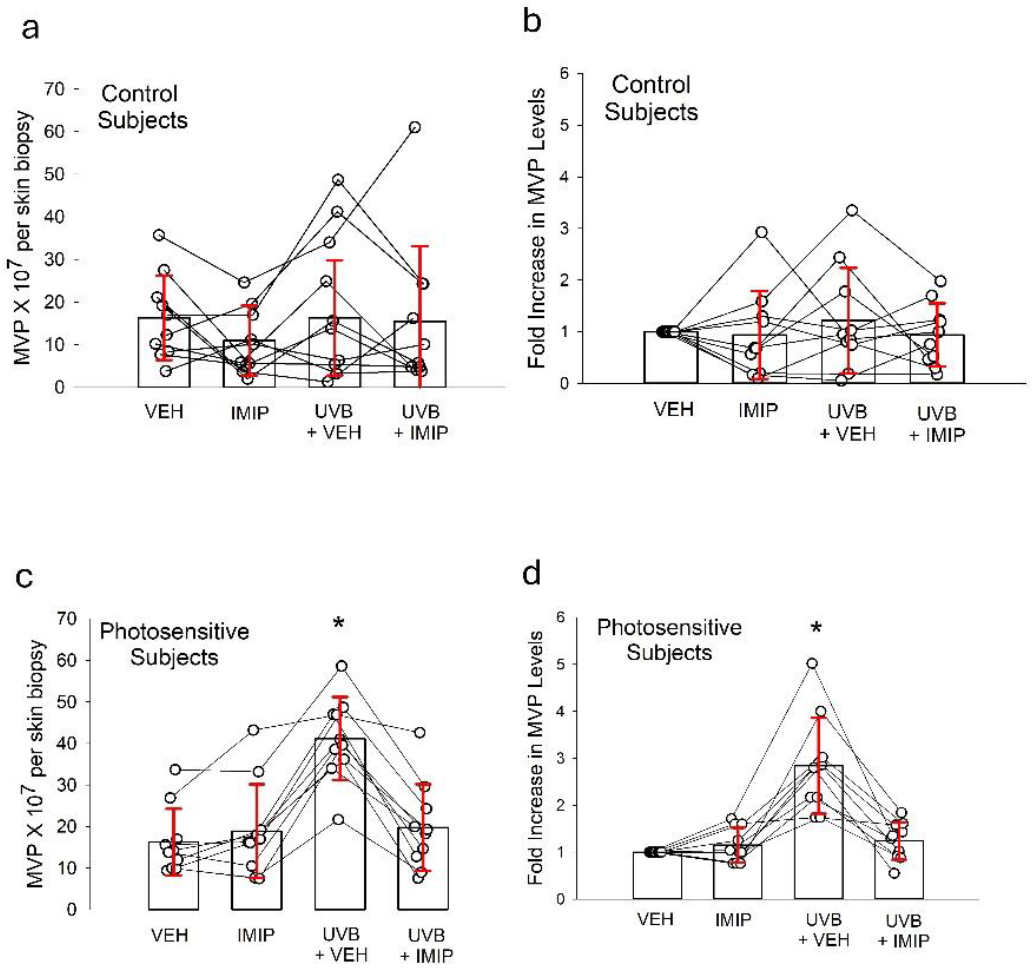
UVB-generated MVP in volar forearm skin of photosensitive versus control subjects and effect of topical imipramine. **a**,**b)** Subjects who did not have a history of photosensitivity (Control Subjects) underwent treatment of two 1 x 1 cm2 areas on volar left forearms with 1000J/m2 UVB, with application of 4% imipramine or vehicle immediately afterwards. On the right volar forearm the subjects were treated with topical imipramine or vehicle. After 4 h, skin biopsies of the four sites (VEH, IMIP, UVB+VEH, UVB+IMIP) were obtained and MVP isolated and quantified. The data are individual values of **a)** absolute MVP numbers per 5 mm skin biopsy or **b)** fold increase in MVP from vehicle-treated skin. **c**,**d)** Similar studies in self-identified photosensitive subjects. Error bars denote the mean ± SD values. Statistically significant *(*P* < 0.05), changes from vehicle values.

Skin erythema is an outcome associated with UVB. Changes in erythema, as measured using a Mexameter, were assessed at four hours following irradiation with UVB of forearm skin treated with vehicle versus imipramine. As depicted in Figure 3, erythema levels increased in both control and photosensitive subjects. Of interest, fold increased levels of erythema were not appreciably different in normal versus photosensitive subjects. However, treatment with imipramine inhibited erythema levels in a statistically significant manner selectively in the photosensitive subjects.

**Figure 3.**
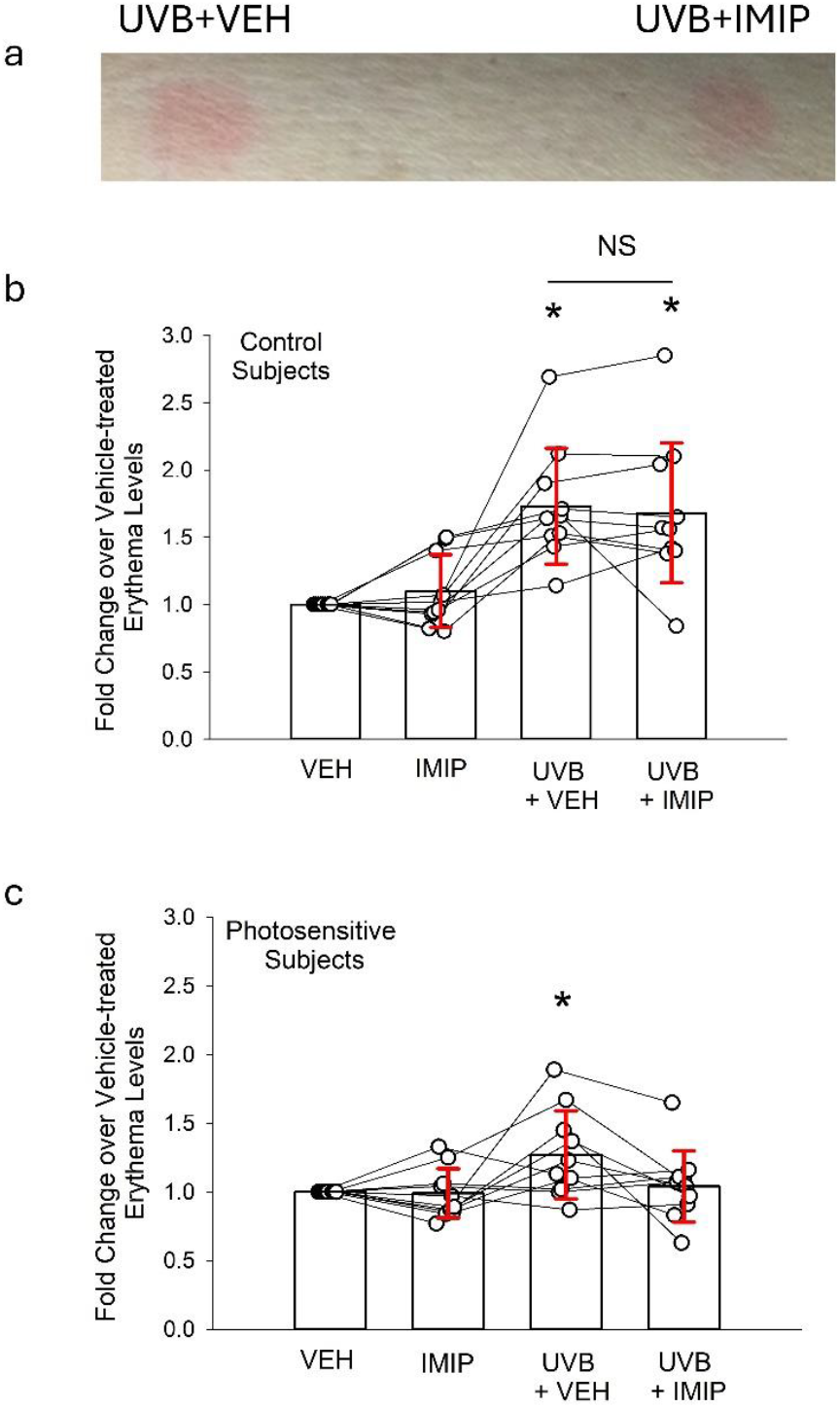
UVB-induced erythema in volar forearm skin of photosensitive versus control subjects and effect of topical imipramine. Subjects who did not have a history of photosensitivity (Control Subjects) or Photosensitive Subjects underwent the identical protocol as outlined in Figure 2. Erythema values were obtained pre- and four hours post-treatment using a mexameter and expressed as fold change from vehicle-treated values. a) Typical example of UVB-induced erythema values at 4 h in a photosensitive subject. b) Data for Control Subjects; c) Data for Photosensitive Subjects. The error bars are mean ± SD fold increase from vehicle-treated values. Statistically significant *(*P* < 0.05) changes; N.S., not statistically significant.

As depicted in Figure 1b, non-invasive studies were conducted on this cohort testing the long-term effects of topical imipramine versus placebo at 24 and 72 hours by measuring erythema levels in response to a range (0-800 J/m2) of UVB fluences which did not reveal statistically significant differences (see Figure 4a,b for 24 h examples; individual data not shown). Nor did the presence/absence of FIASM affect the minimal erythema doses (MED) at 24 h or any of the erythema measurements at 72 h. Examination of the complete subject dataset (**Table I**) revealed an inverse relationship between Minimal Erythema Dose (MED 24 h) and MVP fold increase following UVB exposure (Pearson’s r = −0.55, R^2^ = 0.30, p = 0.013). Participants with low MED values (100–200 J/m^2^, n = 7) showed markedly higher MVP fold increases than those with high MED values (400–600 J/m^2^, n = 6; 3.11 ± 1.17 vs. 1.40 ± 0.78, t(11) = 3.40, p = 0.006, Cohen’s d = 1.4). By contrast, when the same participants were classified according to their self-reported photosensitivity, the photosensitive group released only 0.66 SD more MVP than controls, a difference that was not statistically significant (t(15) = 1.40, p = 0.18). This discrepancy highlights the importance of objective MED measurements over subjective assessments in identifying biologically meaningful differences in UVB responses.

**Figure 4.**
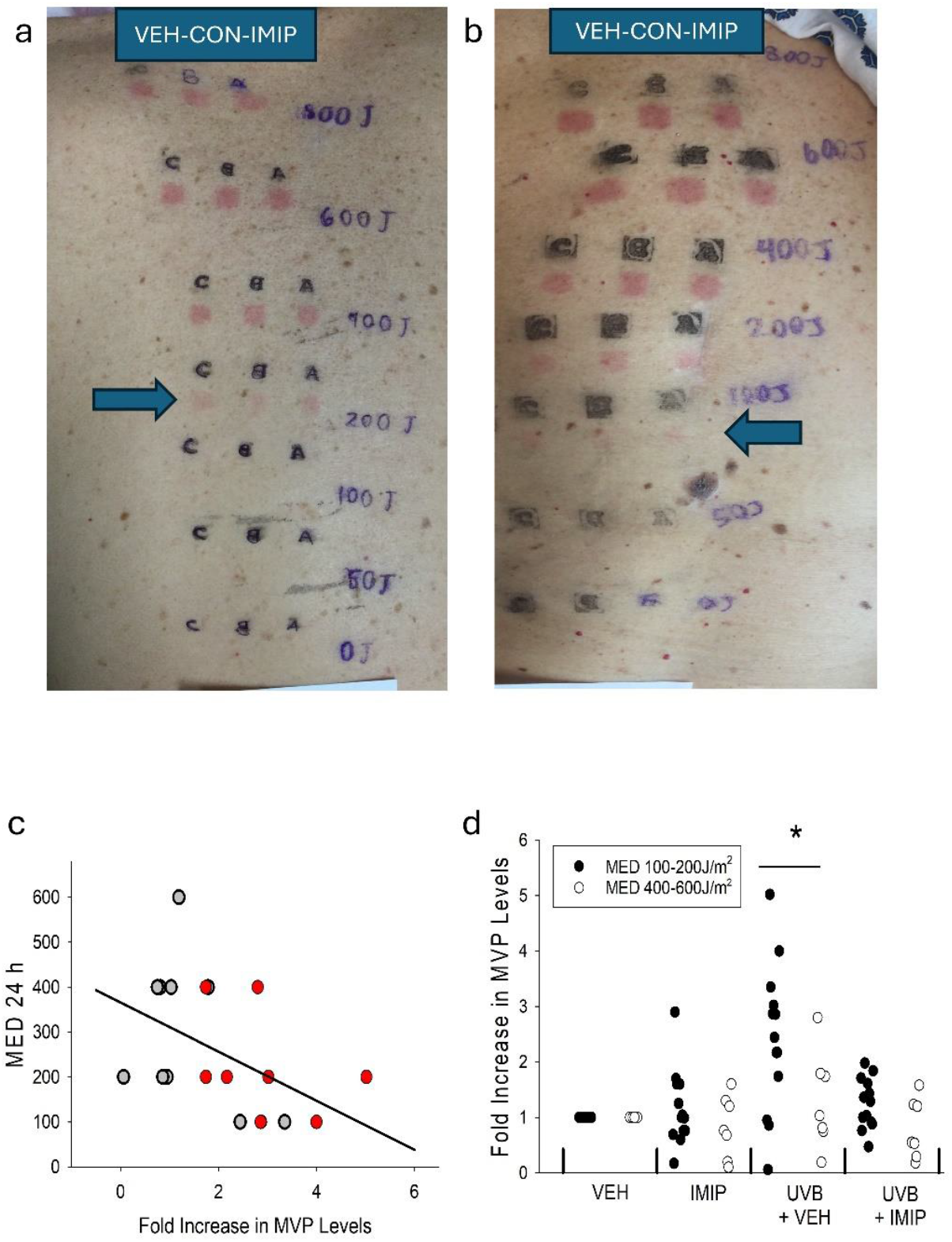
Examples of erythema reactions at 24 h demonstrating lack of effectiveness of topical imipramine at this time following a single post-UVB treatment and comparing MED 24h versus fold MVP increases at 4 hours. Subjects underwent irradiation of back skin with various fluences (0, 50, 100, 200, 400, 600, 800J/m2) of UVB and immediately afterwards were treated with 4% imipramine, vehicle, or no treatment (control). **a**,**b)**. Examples of erythema reactions at 24 h post-UVB revealing no discernable changes in erythema levels between the treatment groups. Blue arrows represent MED 24 **a)** 200J/m2; **b)** 100 J/m2. **c)**. Scatter plot showing significant inverse correlation between MED 24 h values and MVP fold increase (Pearson’s r = −0.55, R^2^ = 0.30, p = 0.013). **d)**. Group comparison of MVP fold increases between objectively defined photosensitive (MED 100–200 J/m^2^, n = 7) and less sensitive (MED 400–600 J/m^2^, n = 6) participants, showing a significant difference (t(11) = 3.40, p = 0.006, Cohen’s d = 1.4). Data presented as mean ± SD with individual data points overlaid. Fold Increase in MVP levels at 4 h versus MED 24 values for the subjects (grey-control; red-self-identified photosensitive). Pearson correlation coefficient (**r**):-0.55; (**R2**):0.30; *p = 0*.*013*. Statistically significant *(*P* < 0.05) changes.

The results of the current studies provide some evidence that implicate cutaneous MVP release in the subsequent acute UVB responses in human subjects. However, the preclinical data from previously published murine studies using photosensitive mice examined at 4 h post UVB is far more compelling (Christian et al., 2024; Corbin et al., 2023). It should be noted that in the XPA deficient model of photosensitivity, both solar simulated light and UVB sources generated equal effects as measured by erythema, cytokine levels and MVP (Christian et al., 2024). This finding suggests that UVB wavelengths would be appropriate for these studies. The exact cell type that is shedding the MVPs in response to UVB is likely the keratinocyte. This is supported by the nature of UVB in that the depth of penetration makes the keratinocyte-rich epidermis the major target. Moreover, previous studies examining both mice and human volunteers undergoing medical phototherapy have reported that the increased MVP found systemically in these populations are positive for the epithelial marker, the calcium-sensing receptor (Liu et al., 2021).

That imipramine appeared to block both the UVB-induced MVP as well as the early erythema levels selectively in photosensitive subjects fits with the hypothesis that these two processes are related. The negative correlation between fold MVP release in skin versus MED 24 noted in all subjects also provides some support for a link between MVP release and skin erythema responses. One possible explanation for the lack of responsiveness of topical imipramine at time points longer than 4 h was that the concentration and/or bioavailability of the FIASM is not optimal. Current studies are underway testing higher strengths (10%) of both imipramine and amitriptyline in photosensitive rosacea patients.

It should be noted that previous studies performed on non-photosensitive subjects before and following supplementation with 7 days of oral antioxidant vitamins (2 g per day of vitamin C and 1000 IU vitamin E per day) blocked the UVB-generated MVP but not the erythema responses (McGlone et al., 2022). Murine studies using wild-type and XPA-deficient mice have also revealed that aSMase inhibition with FIASM or genetic knock-out mice selectively inhibits the 4 h post-UVB erythema responses on the latter photosensitive mice (Christian et al., 2024). The exact cause for UVB-induced erythema is unclear, though eicosanoids and nitric oxide have been implicated (Deliconstantinos et al., 1995; Rhodes et al., 2009). Possible interpretations of our findings include that MVP generation might not be directly involved in erythema responses, but serve as a marker for UVB responses. As MVP contents reflect the stimulated keratinocyte, it is possible that there could be differences in biologic agents found in UVB-induced MVP in normal versus photosensitive subjects. The present studies did not examine skin cytokine levels unlike previously published XPA deficient murine studies (Christian et al., 2023). Current studies are ongoing to define potential differences in the constituents of UVB-generated MVP derived from control versus photosensitive models to assess changes of particle contents which could possibly explain the function(s) of UVB-generated MVP as well as differences associated with photosensitivity.

There are multiple limitations of the current pilot studies. First, the differentiation of both photosensitive and control subjects were based upon self-identification. The lack of statistically significant differences in MED 24 values in these two groups (Table I and Figure 4c) is suggestive that these are not two unique populations. Yet segregation of these subjects based upon MED 100-200J/m2 versus MED 400-600J/m2 did reveal greater fold changes in MVP release in the MED 100-200J/m2 group (Figure 4d). Second, only a single relatively high fluence of UVB (1000 J/m2; 1.7-10 times the MED 24 of the population studied) was employed for the acute 4 h erythema and MVP release data, which might not have been able to adequately differentiate between normal and photosensitive subjects. It is also possible that MVP could be involved selectively in high UVB fluences. Additionally, as noted in Table I, many of the photosensitive subjects were currently treated with systemic agents that would likely tend to blunt UVB reactions. This is a critical item as it would not be ethical to withhold therapies in this population for experimental purposes. Of interest, we found that one of our subjects in the photosensitive group was taking duloxetine, a known FIASM (Kornhuber et al., 2008). As noted in Table I, the data from this subject was not used in these studies but appeared to not respond to UVB with MVP release. The photosensitive subjects were also older than the control subjects, which is relevant as UVB-induced erythema responses (but not MED values) have been reported to be greater in older populations (Gloor and Scherotzke, 2002).

In summary, the current pilot studies sought to provide support for the notion that MVP could play a role in abnormal UVB responses. The ability of an inhibitor of the MVP-generating enzyme aSMase applied post irradiation to attenuate the MVP release as well as blunt the early (4 h) erythema responses and the correlation between MVP release and MED 24 support this hypothesis. Yet, imipramine treatment did not affect erythema responses at later time points with lower fluences and did not change the MED 24 levels. Finding that aSMase inhibitors (e.g., FIASMs) could potentially play a beneficial role in treating photosensitivity could provide a needed therapeutic target. These agents such as tricyclic antidepressants like imipramine are commonly used orally or topically to treat pain with a favorable safety profile (Leppert et al., 2018; Zin et al., 2008). No untoward effects of topical imipramine were noted in these studies. Further studies in this area are needed, especially to ensure that targeting aSMase would not result in long term consequences such as increased photocarcinogenesis by limiting acute UVB effects.

Of interest, a recent multicenter double-blinded placebo-controlled clinical trial reported that oral paroxetine was effective in treating patients with rosacea who exhibited moderate-to-severe erythema (Wang et al., 2023). Though the authors of this important study posited that paroxetine’s effectiveness was due to its well-known ability to act as a selective serotonin reuptake inhibitor with direct effects on the vasculature, paroxetine is also a known FIASM (Kornhuber et al., 2008; Kornhuber et al., 2010). Inasmuch as many patients with rosacea note that UV light is a known trigger (Fisher et al., 2023; McCoy, 2020), it is possible that a component of paroxetine’s effectiveness in this clinical trial could be due to blockade of MVP release. The current studies provide possible impetus for future investigations testing FIASMs in photosensitive disorders to include rosacea which could result in novel therapeutics.

## Materials and Methods

### Human studies

All studies involving humans were approved by the Wright State University Institutional Review Board, Dayton Ohio and followed the Declaration of Helsinki Principles. This clinical trial is registered at ClinTrials.gov (NCT04520217). Volunteers provided written informed consent before enrollment. To assess potential photosensitive subjects, we employed a brief screening questionnaire (Corbin et al., 2023). All subjects were not taking any medications that act as FIASMs nor were taking any medications that are potentially photosensitive nor on any antioxidant vitamins for the past month (McGlone et al., 2022). Photosensitive subjects were allowed to remain on their medications which were designed to treat their underlying disorder (e.g., hydroxychloroquine).

For the Acute 4 h studies, subjects were irradiated with 1000 J/m2 UVB on two 1 x 1 cm2 areas of left volar forearm skin at least 8 cm away from each other using our Philips F20T12/UVB lamp source (Somerset, NJ, USA) using Kodacel filter to remove UVC (McClone et al., 2022). The intensity of the UVB source was measured before each experiment using an IL1700 radiometer and a SED240 UVB detector (International Light, Newburyport, MA, USA) at a distance of 8 cm from the UVB source. Immediately after the UVB irradiation, one area on the left forearm was treated in a 2.5 x 2.5 cm2 area with 0.1 mL of 4% imipramine or 90% polyethylene glycol: 10% DMSO vehicle alone. These solutions were formulated by The Compounding Pharmacy (Huber Heights, Ohio) and were blinded to the investigator applying the agents and the subject. An unblinded member of our clinical trial staff did not have any contact with the subjects. On the right volar forearms, the agents were applied to normal (unirradiated) skin to serve as controls. After 4 hours, erythema measurements were obtained from UVB- and contralateral sham-treated volar forearm skin using a Mexameter, and 5 mm punch biopsies were obtained from all four sites using xylocaine as intradermal anesthetic (McGlone at al., 2022; Liu et al., 2021).

These studies also tested the effects of topical vehicle vs imipramine post-treatment on a series of UVB fluences on the back with the goal of examining later time points. As depicted in Figure 1c, subjects underwent treatment of UVB using fluences of 0 (sham), 50, 100, 200, 400, 600 and 800 J/m2 UVB and immediately afterwards the 1 x 1 cm2 areas were treated with 0.1 mL of either 4% imipramine or vehicle, or left untreated (control). At 24 and 72 hours post UVB, the areas were photographed and erythema measured in blinded fashion using a Mexameter. The MED 24h values were determined by visualization of photographs in by a member of the investigative team who was blinded as to subject details.

### MVP isolation and quantitation

MVP were isolated from skin biopsies as previously reported (McGlone at al., 2022; Liu et al., 2021). For skin tissues, 5 mm punch biopsies were taken, fat was removed with surgical scissors, and the tissues weighed. To the tissue, 0.5 mL of collagenase/dispase (final concentration 5 mg/mL) was added, and then the tissue was finely minced with surgical scissors. The tube containing the tissue was placed in a shaking water bath overnight at 37 °C. The next day, 1 mL of filtered PBS was added and centrifuged initially at 2,000 x g for 10 min at 4 °C. The supernatant was centrifuged at 20,000 x g for 10 min at 4 °C. If the supernatant is not clear, then this step is repeated. Finally, supernatants were centrifuged at 20,000 x g for 70 min at 4 °C. The supernatant was discarded, and the tube was placed upside down to dry for 5 min. The pellet was resuspended with 100 μL of filtered PBS. The samples were either analyzed immediately or kept at 4 °C for less than 1 week, 20 °C for one month, or 80 °C for longer time periods. The concentration of the MVP was determined by using a NanoSight NS300 instrument (NanoSight Ltd, Malvern Instruments, Malvern, UK). Three 30-second videos of each sample were recorded and analyzed with NTA software version 3.0 to determine the concentration and size of measured particles with corresponding standard error. This methodology has been validated using transmission electron microscopy and immunoblotting with selective markers to confirm the identity of MVP fraction (Liu et al., 2021).

### Measurement of erythema levels

The Mexameter MX 18 probe (Courage-Kasaka, San Francisco, CA) was used to measure erythema from the skin as previously described (McGlone et al., 2022; Christian et al., 2024). Three readings were collected both before and after the 4-hour incubation period. By comparing the differences to the baseline value for each site, differences were determined as a measure of change in erythema.

### Statistics

Data normality was assessed using the Shapiro-Wilk test, with all variables demonstrating normal distribution (W > 0.93, p > 0.50). Comparisons between groups were performed using Welch’s two-sample t-test for normally distributed data and Mann-Whitney U test for non-parametric data. Correlations between MED and MVP metrics were evaluated using Pearson’s product-moment correlation coefficient, with Spearman’s rho calculated as a sensitivity measure. For multiple group comparisons, one-way or two-way ANOVA was conducted, followed by Tukey’s post hoc test to control for multiple comparisons. Within-subject analyses utilized paired t-tests, while subgroup comparisons based on photosensitivity stratification employed unpaired two-tailed t-tests. Continuous variables are presented as mean ± standard deviation unless otherwise indicated. Effect sizes are reported as Cohen’s d where applicable. Statistical significance was defined as p < 0.05, and all analyses were conducted using Python 3.11 (SciPy 1.12) and GraphPad Prism 9.0 (GraphPad Software, San Diego, CA).

## Data Availability

All data produced in the present study are available upon reasonable request to the authors

## Ethics Statement

The authors state CAR and JBT along with Wright State University and the US Veterans Administration have obtained a preliminary patent for use of topical acid sphingomyelinase inhibitors in photosensitive disorders.

## Acknowledgments

This research was supported in part by National Institutes of Health grants R01 HL062996 (J.B.T.), R01 ES031087 (J.B.T., M.G.K.), R01GM130583 (M.G.K.) and Veteran’s Administration Merit Awards 5I01BX000853 (J.B.T.) and I01CX002241 (M.G.K.).

